# A geospatial examination of specialist care accessibility and impact on health outcomes for patients with acute traumatic spinal cord injury in NSW, Australia: a whole population record linkage study

**DOI:** 10.1101/2020.07.21.20158899

**Authors:** Lisa Nicole Sharwood, Bharat Phani Vaikuntam, Christiana Cheng, Vanessa Noonan, Anthony Joseph, Jonathon Ball, Ralph Stanford, Mei-Ruu Kok, David Whyatt, Samuel Withers, James Walter Middleton

**Affiliations:** University of Sydney, Faculty of Medicine and Health, NSW, Australia; University of NSW, BlackDog Institute, NSW, Australia; University of Technology Sydney, Faculty of Engineering, NSW, Australia; Monash University, Dept. Epidemiology & Preventive Medicine, VIC, Australia; Cure & Care Programs, Praxis Spinal Cord Institute, Vancouver, Canada; Research & Best Practice Implementation, Praxis Spinal Cord Institute, Vancouver, Canada; Royal North Shore Hospital, NSW, Australia Clinical Associate Professor, University of Sydney, NSW, Australia; Prince of Wales Hospital, NSW, Australia; University of Western Australia, WA, Australia; University of Western Australia, Faculty of Health and Medical Sciences, WA, Australia; Australian Institute of Robotic Orthopaedics, WA, Australia; Rehabilitation Medicine, University of Sydney, NSW, Australia; State-wide Spinal Cord Service, NSW, Australia

## Abstract

**Background:** Timely treatment is essential for achieving optimal outcomes after traumatic spinal cord injury (TSCI), and expeditious transfer to a specialist spinal cord injury unit (SCIU) is recommended within 24 hours from injury. Previous research in New South Wales (NSW) found only 57% of TSCI patients were admitted to SCIU for acute post-injury care; 73% transferred within 24 hours from injury.

**Methods:** This record linkage study included administrative pre-hospital, admissions and costs data for all patients aged ≥16 years with incident TSCI in NSW (2013-2016). Its aim was to examine potential geographical disparities in access to specialist care following TSCI using geospatial methods, and to better understand the impact of post-injury care pathways on patient outcomes.

**Results:** Of 316 cases with geospatial data, injury location analysis showed that over half (53%, n=168) of all patients were injured within 60 minutes road travel of a SCIU, yet only 28.6% (n=48) were directly transferred to a SCIU. Direct transfers received earlier operative intervention (median (IQR) 12.9(7.9) hours), compared with patients transferred indirectly to SCIU (median (IQR) 19.5(18.9) hours), and had lower risk of complications (OR 3.2 v 1.4, p<0.001).

**Conclusions:** Getting patients with acute TSCI patients to the right place at the right time is dependent on numerous factors; some are still being triaged directly to non-trauma services which delays specialist and surgical care and increases complication risks. More stringent adherence to recommended guidelines would prioritise direct SCIU transfer for patients injured within 60 minutes radius, enabling the benefits of specialised care.

## INTRODUCTION

Traumatic spinal cord injury (TSCI) is a devastating condition with lifelong physical, psychosocial and economic impacts^1-5^. Acute post-injury phase management is time-sensitive, with rapid access to specialist care deemed essential for achieving optimal outcomes. Expeditious transfer to a specialist spinal cord injury unit (SCIU) is recommended within 24 hours following injury^6-8^. Previous research in New South Wales (NSW), Australia^9^ found only 57% of patients with confirmed TSCI admitted to a SCIU for acute care, and of these, only 73% were transferred within 24 hours from injury. Rurality of residence was associated with more timely admission to SCIU in this study^9^, geospatial information system (GIS) data for injury incidents was unable to support this claim.

Specialist health services are often concentrated in high population density areas, though may often serve large geographical areas with substantial, distributed populations. Time and distance consequently challenge equitable access to these services for the whole population. NSW is the most populous Australian state, covering an area of 809,444 km^2^. NSW specialist health services include the State Spinal Cord Injury Service (SSCIS) network, providing care for approximately 5,500 people living with spinal cord injury (2019)^10^. TSCI is a high acuity, resource intensive injury requiring ongoing acute and rehabilitative care; in 2017-18, there were 3,888 re-hospitalisations of people living with spinal cord injury in NSW.

Epidemiological, health services studies are increasingly using geospatial methods^11-13^ to better understand and target both injury prevention and post-injury care^14^. Triage optimisation using geospatial data and scenario modelling has demonstrated significantly reduced time to SCIU admission in a Canadian study^15^. Similar modelling (without geospatial data) showed that optimizing patient-care pathways can achieve significant health system cost reductions from direct in comparison with indirect admissions to SCIU^16^. Studying individuals with acute traumatic brain injury (TBI), Brown et al^12^ found significant disparity in survival rates between rural and urban areas using geospatial analysis. Specifically, in the most rural areas, TBI fatality rates were 13 deaths per 100,000 persons higher than those in the most urban area (95% confidence interval 12.15-13.86; P < 0.001)^12^. A similar analysis across the NSW trauma system demonstrated a significantly higher adjusted mortality rate for patients with any traumatic injury treated in regional services compared with metropolitan major trauma services^17^. Definitive care at a Major Trauma Service (MTS) was associated with a 41% lower likelihood of death compared to definitive care at a Regional Trauma Service (RTS) (OR 0.59 95%CI 0.35-0.97)^17^.

Lack of access to specialist care services, including major trauma management^18^, timely and appropriate surgical interventions within a critical time window and other factors such as increased rates of complications are likely contributing to higher mortality rates and poor long term outcomes for survivors. There is an indication internationally that regardless of prescribed, evidence-based pathways for trauma patients and injury locations within acceptable proximity to specialist services, some acute trauma patients are still being triaged to non-trauma services^19^. Delivery of health services must account for the fact that injuries will occur in a range of geographic locations, from urban to very remote areas. Triaging systems that direct patient pathways through pre-hospital and acute care settings must therefore offer equitable opportunity for optimal outcomes regardless of the incident location. Geospatial analysis for victims of burn injury, as well as bicyclist crash, has been previously conducted in NSW, targeted towards informing public health interventions and aid policy makers plan for service provision^20 21^. The extent to which geospatial variables impact pathways to direct SCIU admission for patients with TSCI in Australia has not been examined.

The aim of this study was to use geospatial methods to investigate the impact of geographical location on pathways and timing of admission to specialist care services for individuals sustaining acute TSCI across the state of NSW, Australia. Analyses will control for relevant variables and assess associations with patient outcomes, including time to surgery and inpatient complications.

The NSW Population Research Ethics Committee (2012/09/420) approved this project.

## MATERIALS AND METHODS

### Setting and acute Health Services

The state of NSW, Australia, has a population of just over 8 million persons^22^ and covers a geographically diverse area > 800,000 km^2^. Around two-thirds of the population reside in the Greater Sydney (suburban) area; the remainder in rural and very remote areas. The state government funded NSW Ambulance Service is the sole emergency medical service, transporting patients via road, fixed wing or helicopter depending on injury severity and geographic location. Road ambulances are not routinely staffed with emergency physicians, whereas some helicopter services are.

Trauma service hospitals are designated as either MTS (equivalent to Level 1 Trauma Service, accredited) or RTS (equivalent to Level 3)^23^; critically ill patients can be taken to one of six strategically located MTS or ten RTS. Approximately 200 additional non-trauma designated hospitals (district/regional/local hospitals) are situated across metropolitan and regional health districts around the state. The SSCIS comprised two specialist SCIUs, both located in metropolitan Sydney. One of these is also a MTS (categorised as SCIU for this study), the other a non-trauma designated hospital; both providing specialist spinal surgical services for TSCI. Hospitals were categorized as trauma hospitals (MTS or RTS), specialist hospitals (SCIU) or non-trauma hospitals (district/local) for the analysis.

Being suspected of having TSCI or evidence of TSCI meets major trauma criteria under the NSW state-wide pre-hospital triage criteria applies; whereby ‘patients meeting major trauma criteria should be transported to the highest level Trauma Centre located within a 60-minute driving radius, which may include bypassing closer non-trauma hospitals’^24^. Recommendation from the SSCIS was that once medically stable, patients with TSCI should be transferred to an SCIU within 24 hours from injury^25^.

### Population-level record linkage

The NSW Admitted Patient Data Collection (APDC) was used to identify and extract TSCI patient records from all separations/discharges from NSW public hospitals, based on specific TSCI-related International Classification of Diseases, 10th version, Australian Modification (ICD-10-AM) diagnosis codes^26^. Probabilistic data linkage was undertaken by the NSW Centre for Health Record Linkage, linking all patients where a TSCI code was either a principal or additional diagnosis, for any separation within the APDC. Included patients were aged 16 years or more, injured between 1 June 2013-30 June 2016, admitted to a NSW hospital and diagnosed as having TSCI using ICD-10AM diagnostic codes^27^. Excluded were those with missing GIS data for these analyses. The first hospital episode and all contiguous care episodes, including nested/non-nested transfers, was deemed the ‘index admission’.

TSCI case records were linked with Emergency Department and NSW Ambulance data. All datasets and variables included are detailed in Appendix 1.

### Outcome variables

Primary outcome was triage pattern to SCIU (direct or indirect admission vs no admission) for acute incident TSCI patients. Direct admissions included those first transferred to SCIUs, indirect admissions were those transferred to SCIU hospitals from all non-SCIU hospitals. The definitive hospital was defined as the final hospital where the patient received their acute care, prior to either rehabilitation admission, discharge home or death, indicated by acute care-type and separation/discharge destination.

Secondary outcomes considered included time to surgery (for operated patients) and incidence of inpatient complications, adjusted for relevant predictor variables including triage pattern to SCIU. Inpatient complications included pressure injuries, urinary tract infections, respiratory infections, deep vein thrombosis, pulmonary embolus and others.

### Geospatial mapping

Python language was used to conduct geospatial analyses, interacting with Google Distance Matrix Application Programming Interfaces (API) to calculate on-road travel distances and times relative to hospitals/SCIUs using injury location GIS co-ordinates. Travel duration was classified as <60 minutes or >60 minutes from SCIU, and/or MTS from injury location, calculated for each patient. Population dispersement data (ABS)^22^ enabled risk of disease mapping per unit-head of population over the study time-period according to Statistical Area (SA) boundaries. Population sizes for different SAs for the years 2012-2015 were calculated by linear interpolation using the 2011 and 2016 populations. Crude rates/100,000 person-years were calculated, and due to the sample size, SA3 determined the most stable representation, using person-years between 1st June 2013-30th June 2016 (Figure 1).

**Figure 1:**
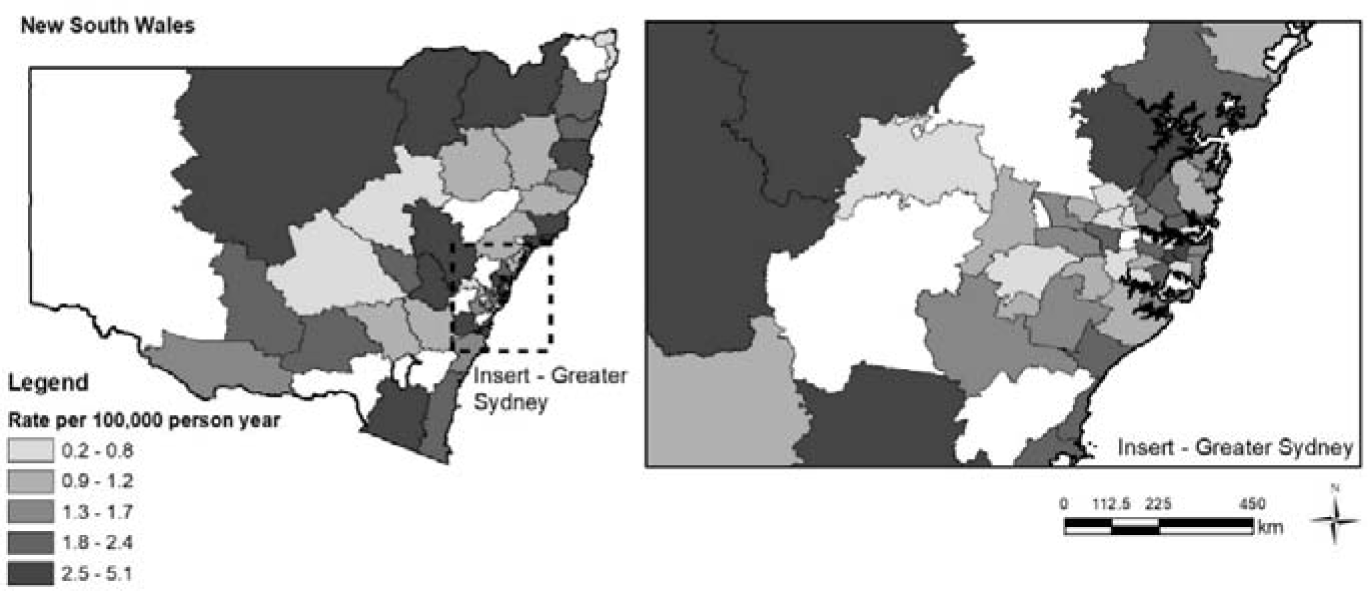
Crude incidence rate per 100,000 person years by SA3.

### Statistical analysis

Descriptive analysis included continuous variables summarised as means (standard deviations) and medians (interquartile range); categorical variables as percentages. Statistically significant differences (<0.05) between groups were tested using t-tests, Wilcoxon Rank Sum or chi-square tests. Multinomial logistic regression modelling examined associations between predictor variables and direct/indirect/no admission to SCIU. Age, gender, injury mechanism, transport mode, presence of multi-trauma, co-morbidity (CCI), geographic distance from incident location relative to SCIU hospital, travel time to SCIU, and injury severity (ICISS score) were included in the regression analysis for the entire study population. Admission times to ‘definitive hospitals’ were used from variables denoting the first admission time to the final (definitive) hospital for the acute care stay. For those patients not transferred, this was also the first hospital to which they were admitted. A multinomial logistic regression model^31^ was assessed using Relative Risk Ratios (RRR), obtained by exponentiating the multinomial logit coefficients, e^coef^. Modelling used the category ‘Direct to SCIU’ as the referent group; the RRR for the groups ‘Indirect to SCIU’ and ‘No SCIU’ indicating risks relative to this referent group. STATA-IC.v.16^32^ was used for all analyses.

## RESULTS

### Demographic and injury profile

During the study period, 534 patients with acute incident TSCI were identified across NSW; 316 (59%) had GIS data for inclusion in geospatial analysis. Injury mechanisms were commonly falls (n=149, 47.2%) and transport crashes (n= 99, 31.3%), with 53% (n=168) sustaining multiple trauma. Over one-third of patients (n=126, 39.9%) had higher injury severity scores (ICISS<0.83)^28^; the median (IQR) ICISS score was 0.839 (0.137) (Table 1); over half of all patients sustained a cervical level injury (n=170, 53.8%).

**Table 1.** Comparing patients with TSCI admitted directly or indirectly to SCIU, or not at all.

| Variable | Direct admit<br>to SCIU<br>(n=85, 26.9%) | Indirect<br>admit to<br>SCIU<br>(n=108, 34.2%) | No acute<br>admit to<br>SCIU<br>(n=123, 38.9%) | Total<br>(n=316) | p-value |
| --- | --- | --- | --- | --- | --- |
|  | **n (%)** | **n (%)** | **n (%)** | **n (%)** |  |
| **Age** years (median (IQR)) | 57.7 (28.9) | 52.7 (37.1) | 60.6 (38.1) | 56.5 (36.3) | 0.05 |
| 16-30 | 17 (20) | 27 (25) | 18 (14.6) | 62 (19.6) |  |
| 31-45 | 12 (14.1) | 20 (18.5) | 17 (13.8) | 49 (15.5) |  |
| 46-60 | 22 (25.9) | 23 (21.3) | 28 (22.8) | 73 (23.1) |  |
| 61-75 | 20 (23.5) | 24 (22.2) | 22 (17.9) | 66 (20.9) |  |
| 76+ | 14 (16.5) | 14 (12.9) | 38 (30.9) | 66 (20.9) |  |
| **Sex** |  |  |  |  | 0.10 |
| Female | 16 (18.8) | 22 (20.4) | 37 (30.1) | 75 (23.7) |  |
| Male | 69 (81.2) | 86 (79.6) | 86 (69.9) | 241 (76.3) |  |
| **Injury mechanism** |  |  |  |  | 0.15 |
| Fall | 43 (50.6) | 45 (41.7) | 61 (49.6) | 149 (47.2) |  |
| Transport | 32 (37.6) | 35 (32.4) | 32 (26.0) | 99 (31.3) |  |
| Self-harm/Assault | 1 (1.2) | 8 (7.4) | 7 (5.7) | 16 (5.1) |  |
| Other<sup>‡</sup> | 9 (10.6) | 20 (18.5) | 23 (18.7) | 52 (16.5) |  |
| **Incident GPS** |  |  |  |  | 0.74 |
| ≤ 60min to SCIU | 48 (56.4) | 55 (50.9) | 65 (52.8) | 168 (53.2) |  |
| > 60min to SCIU | 37 (43.5) | 53 (49.1) | 58 (47.1) | 148 (46.8) |  |
| **SEIFA Quintiles** |  |  |  |  | 0.002 |
| 1 (lowest) | 8 (9.4) | 21 (19.4) | 15 (12.2) | 44 (13.2) |  |
| 2 | 14 (16.5) | 24 (22.2) | 29 (23.6) | 67 (21.2) |  |
| 3 | 11 (12.9) | 23 (21.3) | 37 (30.1) | 71 (22.5) |  |
| 4 | 15 (17.6) | 11 (10.2) | 17 (13.8) | 43 (13.6) |  |
| 5 (highest) | 35 (41.2) | 28 (25.9) | 20 (16.3) | 83 (26.3) |  |
| unknown | 2 (2.3) | 1 (0.9) | 5 (4.1) | 8 (2.5) |  |
| **Charlson (CCI)** |  |  |  |  | 0.84 |
| 0 | 60 (70.6) | 77 (71.3) | 82 (66.7) | 219 (69.3) |  |
| 1 | 14 (16.5) | 14 (12.9) | 19 (15.4) | 47 (14.9) |  |
| 2+ | 11 (12.9) | 17 (15.7) | 22 (17.9) | 50 (15.8) |  |
| **Multiple Trauma** |  |  |  |  | 0.003 |
| No | 33 (38.8) | 45 (41.7) | 70 (56.9) | 148 (46.8) |  |
| 1 other injury | 29 (34.1) | 20 (18.5) | 23 (18.7) | 72 (22.8) |  |
| ≥2 other injuries | 23 (27.1) | 43 (39.8) | 30 (24.4) | 96 (30.4) |  |
| **ICISS Score** |  |  |  |  | <0.001 |
| < 0.7 (Most severe) | 9 (10.6) | 5 (4.6) | 11 (8.9) | 25 (7.9) |  |
| 0.7 to <0.83 | 48 (56.5) | 26 (24.1) | 27 (21.9) | 101 (31.9) |  |
| 0.83 to <0.89 | 17 (20) | 29 (26.8) | 23 (18.7) | 69 (21.8) |  |
| 0.89 to <0.95 | 8 (9.4) | 21 (19.4) | 29 (23.6) | 58 (18.4) |  |
| 0.95 to 1.00 | 3 (3.5) | 27 (25) | 33 (26.8) | 63 (19.9) |  |
| **Highest Level of Injury** |  |  |  |  | <0.001 |
| Cervical | 54 (63.5) | 66 (61.1) | 50 (40.6) | 170 (53.8) |  |
| Thoracic | 22 (25.9) | 27 (25) | 34 (27.6) | 83 (26.3) |  |
| Lumbar | 9 (10.6) | 15 (13.9) | 39 (31.7) | 63 (19.9) |  |
| **Extent of Injury** |  |  |  |  | <0.001 |
| Complete | 24 (28.2) | 23 (21.3) | 5 (4.1) | 52 (16.5) |  |
| Incomplete | 43 (50.6) | 57 (52.8) | 39 (31.7) | 139 (43.9) |  |
| Conus /cauda equina | 11 (12.9) | 17 (15.7) | 38 (30.9) | 66 (20.9) |  |
| Unspecified | 7 (8.2) | 11 (10.2) | 41 (33.3) | 59 (18.7) |  |
| **Mode of Arrival** |  |  |  |  | <0.001 |
| Ambulance | 56 (65.9) | 99 (91.7) | 115 (93.5) | 270 (85.4) |  |
| Air ambulance | 18 (21.2) | 5 (4.6) | 3 (2.4) | 26 (8.2) |  |
| Others | 11 (12.9) | 4 (3.7) | 5 (4.1) | 20 (6.3) |  |

‡Other included diving, horse-related incidents or not specified
\*SEIFA – Socio-Economic Indexes for Areas <sup>30</sup>

Table 1 compares baseline demographic and injury epidemiology profile for patients admitted directly, indirectly or not at all to SCIU. Patients were more likely to experience direct transfer to a SCIU without comorbid trauma (p<0.01) but higher ICISS (p<0.001), cervical injury (p<0.01), and transferred by air-ambulance (p<0.01). Indirect transfer to SCIU was more likely with two or more additional traumatic injuries (p<0.01) or incomplete injury (p<0.01). Patients not admitted to SCIU at all were older (p=0.05) with lower levels of injury (p<0.01).

### Case rates across the state of NSW

GIS location of all cases were mapped, and crude rates calculated per 100,000 person-years (Figure 1). Plotting case distribution per SA boundary SA3 was selected as the most stable representation.

### Travel times from incident location

Around one quarter of study patients (n=85, 26.9%) were directly admitted to SCIU from the injury scene, 170 (53.8%) patients went first to an MTS/RTS; leaving 61 patients (19.3%) who went first to local/district hospitals (Table 2). A further 108 (34.2%) were transferred indirectly to SCIU, from one of these other initial destinations, less than 60% transferred within 24 hours from time of injury (n=64, 59.2%). Almost 30% (n=93, 29.4%) of all patients remained in an MTS as their definitive hospital, 69 (74.2%) had been admitted directly from the injury scene. A further 11 (1.8%) were transferred in from an RTS and 13 (13.9%) from local/district hospitals.

**Table 2.** Distances and times to first hospital from injury location GIS, by mode of transport.

| **Hospital Class** | **First Hospital**<br>**n (%)** | **Distance (km) to first hospital**<br>**Median (IQR)** | **Time (mins) to first hospital**<br>**Median (IQR)** | **Definitive Hospital**<br>**n (%)** | **Time (hrs) to definitive hospital<sup>\*\*\*</sup>**<br>**Median (IQR)** |
| --- | --- | --- | --- | --- | --- |
| *Travelled by road ambulance – GIS calculated* |  |  |  |  |  |
| **SCIU** | 67 (23.1) | 20.3 (72.7) | 27.6 (51.6) | 170 (58.6) | 12.6 (39.9) |
| **MTS** | 116 (40.0) | 7.7 (9.9) | 15.6 (11.4) | 90 (31.0) | 23.3 (43.6) |
| **RTS** | 46 (15.8) | 13.4 (29.5) | 18.0 (23.1) | 17 (5.9) | 19.9 (44.4) |
| **Local** | 61 (21.0) | 8.7 (27) | 12.4 (19.5) | 13 (4.5) | 2.9 (0) |
| **Total** | **290** | **9.7 (20.0)** | **16 (17.8)** | **290** | **14.4 (42.9)** |
| *Travelled by air ambulance – direct distances/ recorded times* |  |  |  |  |  |
| **SCIU** | 18 (69.2) | 120 (141.9) | 176 (279) | 23 (88.5) | 8.9 (5.2) |
| **MTS** | 7 (26.9) | 78.4 (34.8) | 153 (33) | 3 (11.5) | N/A |
| **RTS** | 1 (3.8) | 217.3 (0) | 261 (0) | N/A | N/A |
| *Travelled by air ambulance – if had travelled by road (not adjusted for ‘on scene’ time)* |  |  |  |  |  |
| **SCIU** | 18 (69.2) | 151.2 (184.7) | 111.7 (102.9) | N/A | 23 (88.5) |
| **MTS** | 7 (26.9) | 93.4 (58.8) | 68.2 (47.5) | N/A | 3 (11.5) |
| **RTS** | 1 (3.8) | 268.3 (0) | 185.5 (0) | N/A | N/A |
| **Total** | **26** | **122.8 (133.2)** | **168.5 (100.2)** | N/A | **26** |

\*\* Ambulance pick up date/time to definitive hospital admission date/time for those transferred from first hospital

Analysing the GIS data of injury incident locations relative to the proximity to hospital types, we found over half (53%, n=168) of all patients were injured within 60 minutes road travel from an SCIU, yet only 28.6% of these (n=48) were directly transferred to SCIU. Of the remaining patients injured within the 60-minute radius, less than half (48%, n=76) were first transferred to an MTS; 22 (14%) first taken to an RTS. For forty percent (n=125) of all patients the injury incident location was outside the 60 minutes travel radius of any MTS/SCIU, yet 20% went directly to an MTS and 20% to SCIU.

Table 2 shows GIS calculated summary of road travel distances and times calculated from injury incident locations to first hospitals, nearest SCIU and trauma centre by class of first hospital, analysing separately 26 (8.2%) patients who arrived by air-ambulance to their first hospital. For this group, Table 2 shows the actual times and distances (‘direct’ calculations) using hospital admission times and GIS data, also using GIS data to provide the ‘on road’ scenarios, had these patients travelled instead by road.

Differences between the first and second last column above indicate movement between hospital classes, for patients transferred from first to definitive hospitals (n=135, 42.7%). For this transferred group, the median(IQR) time from first to definitive hospital admission was 11.7(38.5) hours; the mean(SD) was 105.7(314.3) hours. Of 193 patients admitted definitively to SCIU, those arriving indirectly, took median(IQR) of 10.5 (38.3) hours, and a mean(SD) of 112.7 (344.4) hours.

For patients *not* taken directly to a SCIU or an MTS but instead taken first to a local/district hospital (n=61), the additional median (IQR) difference in travel times they would have had to make to reach an MTS would have been 79.9 (218.9) minutes; the mean (SD) 133.4 (130) minutes. Almost half (n=28, 46%) could have reached their closest MTS hospital by adding less than 30 minutes to their journey (median(IQR) 14.5(12.7) minutes).

### Time to surgery

Almost two-thirds (n=194, 61.4%) had surgery during acute care admissions; around two-thirds of these had surgery performed at SCIU (n=131, 67.5%). Additionally, 56 (28.9 %) received surgery at an MTS, 7 (3.6%) at an RTS.

Times from ambulance call to surgery were available for 98 (51%) patients; 47 (47.9%) of whom were transferred directly from injury scene to SCIU and had surgery within median (IQR) of 12.9 (7.9) hours. This was more rapid than for patients transferred indirectly to SCIU (from MTS/RTS/local hospitals), who had surgery within median (IQR) of 19.5 (18.9) hours. Patients transferred directly from injury location to MTS and who surgery there, were operated within a median (IQR) of 21.8 (115.2) hours.

Comparing times to admission to hospitals where operations took place for the surgery group of 194 patients, Table 3 compares times to surgical hospital admission for the 98 for whom operation times were available with times to surgical hospital admission for the other 96 patients, as well as times to surgery where available. Inter-hospital transfers lengthened times to surgery.

**Table 3:** First hospital, location of surgery and timing to surgery (median [IQR]) post injury.

| **Hospital Class** | **As First hospital**<br>**n (%)** | **As Surgical Hospital**<br>**n (%)** | **Time to surgery**<br>**median (IQR)**<br>**(by surgical hospital class)** | **Time (hrs) to surgical hospital admission\***<br>**median (IQR)** |
| --- | --- | --- | --- | --- |
| ***Time to surgery available (n=98)*** |  |  |  |  |
| **SCIU** | 47 (47.9) | 90 (91.8) | 15.2 (10.9) | 6.7 (7.3) |
| **MTS** | 31 (31.6) | 7 (7.1) | 21.8 (23.3) | 1.4 (0.5) |
| **RTS** | 8 (8.2) | 1 (1.0) | 12.7 (0) | 1.4 (0) |
| **Local** | 12 (12.2) | 0 | NA | NA |
| **Total** | 98 | 98 | 15.2 (11.2) | 6.1 (7.2) |
| *Time to surgery not available (n=96)* |  |  |  |  |
| **SCIU** | 18 (18.7) | 41 (42.7) | NA | 8.1 (9.4) |
| **MTS** | 45 (46.9) | 49 (51.0) | NA | 1.4 (8.5) |
| **RTS** | 20 (20.8) | 6 (6.3) | NA | 1.0 (0.8) |
| **Local** | 13 (13.5) | 0 | NA | 0 |
| **Total** | 194 | 194 | NA | 3.9 (10.2) |

\*from ambulance pick up time

### Factors influencing triage pattern to SCIU

The multinomial logistic regression model (Table 4) displays the RRR of various exposure variables to triage patterns; with direct SCIU admission as the base outcome category to no SCIU admission, then indirect SCIU admission. Where the nearest SCIU was within 60 minutes road travel from the injury incident the likelihood of no SCIU admission was significantly reduced (RRR 0.281, p=0.003, 95% CI 0.123-0.646). The relative risk of indirect SCIU transfer was somewhat lower (RRR 0.511) than no SCIU. Patients with two or more concurrent traumatic injuries were 5.2 times more likely to experience indirect (rather than direct) transfer to SCIU (RRR 5.18, p=0.002). Patients with lower injury severity (ICISS 0.95-1.00) were around 10 times more likely to have no admission to SCIU than direct transfer compared to patients with higher injury severity (ICISS< 0.7), (RRR 10.1, p=0.03, 95% CI 1.194-85.885). Patients with complete or incomplete TSCI were significantly less likely to experience no SCIU admission in relation to patients without any completeness of injury coded. Each year older for patients with acute TSCI increased the likelihood of not being admitted to a SCIU at all in the acute admission by 2% per year (p=0.032). Table 4 displays the regression estimates for the entire cohort.

**Table 4:** Multinomial regression model – relative risk of exposure variables comparing Indirect and No transfer to SCIU, with reference of direct admission to SCIU

| **Triage Patterns** | **RRR\*** | **Std. Err** | **P-value** | **95% Confidence Interval** |
| --- | --- | --- | --- | --- |
| **Direct SCIU** | **(as base outcome for No SCIU)** |  |  |  |
| **No SCIU** |  |  |  |  |
| age (years) | 1.02 | 0.01 | 0.034 | 1.002 - 1.039 |
| SCIU > 60 min | 0.281 | 0.119 | 0.003 | 0.123 - 0.646 |
| Female sex | 0.959 | 0.44 | 0.928 | 0.391 - 2.355 |
| ***Injury Mechanism*** |  |  |  |  |
| Other (reference) |  |  |  |  |
| Falls | 0.629 | 0.343 | 0.388 | 0.21 - 1.83 |
| Transport | 0.559 | 0.343 | 0.344 | 0.17 - 1.86 |
| Self-harm /Assault | 11.449 | 16.289 | 0.087 | 0.70 - 186.13 |
| ***Injury level*** |  |  |  |  |
| Cervical (reference) |  |  |  |  |
| Thoracic | 1.838 | 0.983 | 0.255 | 0.64 - 5.24 |
| Lumbar | 3.917 | 5.142 | 0.298 | 0.29 - 51.33 |
| ***Extent of spinal cord Injury*** |  |  |  |  |
| Not coded (reference) |  |  |  |  |
| Complete | 0.061 | 0.044 | <0.001 | 0.015 - 0.25 |
| Incomplete | 0.288 | 0.159 | 0.024 | 0.09 - 0.85 |
| Conus/cauda-equina | 0.108 | 0.136 | 0.077 | 0.01 - 1.27 |
| ***Arrival Mode*** |  |  |  |  |
| Missing (reference) |  |  |  |  |
| Road Ambulance | 8.139 | 6.273 | 0.007 | 1.79 - 36.86 |
| Air ambulance | 0.61 | 0.606 | 0.619 | 0.09 - 4.27 |
| ***ICISS Score*** |  |  |  |  |
| <0.7 (base) |  |  |  |  |
| 0.7 to <0.83 | 0.696 | 0.471 | 0.592 | 0.18 - 2.62 |
| 0.83 to <0.89 | 1.558 | 1.154 | 0.55 | 0.36 - 6.66 |
| 0.89 to <0.95 | 2.275 | 1.929 | 0.332 | 0.43 - 11.99 |
| 0.95 to 1.00 | 10.125 | 11.045 | 0.034 | 1.19 - 85.88 |
| ***Multiple trauma*** |  |  |  |  |
| 0 (base) |  |  |  |  |
| One additional injury | 0.35 | 0.158 | 0.02 | 0.15 - 0.85 |
| Two or more | 0.942 | 0.503 | 0.911 | 0.33 - 2.68 |
| **Direct SCIU** | **(as base outcome for Indirect SCIU)** |  |  |  |
| **Indirect SCIU** |  |  |  |  |
| Age (years) | 0.998 | 0.01 | 0.84 | 0.98 - 1.02 |
| SCIU > 60 min | 0.511 | 0.22 | 0.11 | 0.22 - 1.17 |
| Female sex | 0.96 | 0.45 | 0.93 | 0.38 - 2.43 |
| ***Injury Mechanism*** |  |  |  |  |
| Other (reference) |  |  |  |  |
| Falls | 0.51 | 0.28 | 0.22 | 0.17 - 1.49 |
| Transport | 0.56 | 0.34 | 0.34 | 0.17 - 1.84 |
| Self-harm/Assault | 9.02 | 12.77 | 0.12 | 0.56 - 144.74 |
| ***Injury level*** |  |  |  |  |
| Cervical (reference) |  |  |  |  |
| Thoracic | 0.61 | 0.32 | 0.35 | 0.22 - 1.69 |
| Lumbar | 0.23 | 0.27 | 0.22 | 0.02 - 2.48 |
| ***Extent of Injury*** |  |  |  |  |
| Missing (reference) |  |  |  |  |
| Complete | 1.774 | 1.21 | 0.40 | 0.46 - 6.77 |
| Incomplete | 1.625 | 1.03 | 0.44 | 0.47 - 5.62 |
| Conus/cauda-equina | 0.502 | 0.59 | 0.56 | 0.05 - 5.01 |
| ***Arrival Mode*** |  |  |  |  |
| Other (reference) |  |  |  |  |
| Road ambulance | 20.148 | 18.13 | 0.001 | 3.45 - 117.52 |
| Air ambulance | 1.768 | 1.84 | 0.58 | 0.23 - 13.55 |
| ***ICISS Score*** |  |  |  |  |
| <0.7 (reference) |  |  |  |  |
| 0.7 to <0.83 | 2.04 | 1.52 | 0.34 | 0.47 - 8.79 |
| 0.83 to <0.89 | 9.98 | 8.14 | 0.005 | 2.02 - 49.38 |
| 0.89 to <0.95 | 18.69 | 17.09 | 0.001 | 3.11 - 112.25 |
| 0.95 to 1.00 | 33.87 | 39.64 | <0.001 | 24.14 - 58.76 |
| ***Multiple trauma*** |  |  |  |  |
| 0 (reference) |  |  |  |  |
| One additional injury | 0.65 | 0.29 | 0.35 | 0.26 - 1.59 |
| Two or more | 5.18 | 2.77 | 0.002 | 1.82 - 14.75 |

\*Relative Risk Ratios

### Triage patterns influencing complication risk

The potential impact of triage patterns on the risk of experiencing inpatient complications for patients with acute TSCI was assessed and is shown in Table 5. Patients experiencing indirect transfer to SCIU had an increased risk of developing inpatient hospital complications compared with patients transferred directly to SCIU. Complete TSCI, additional comorbidities (CCI) and older age also increased the risks of developing inpatient complications.

**Table 5:**
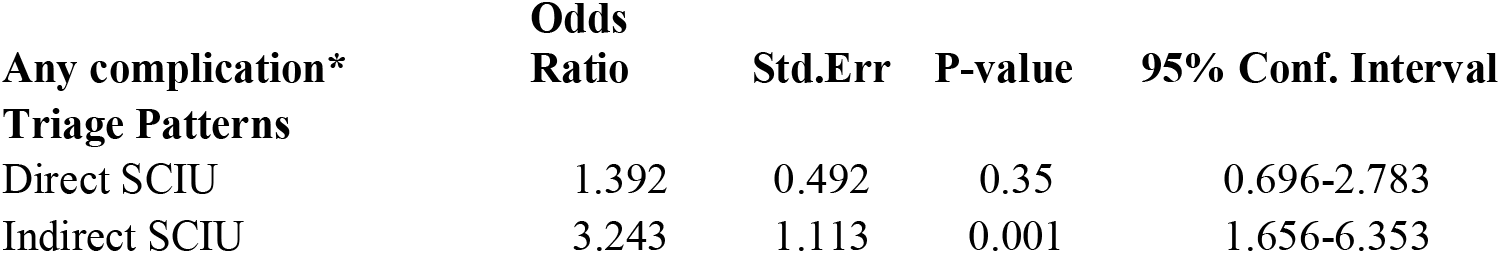
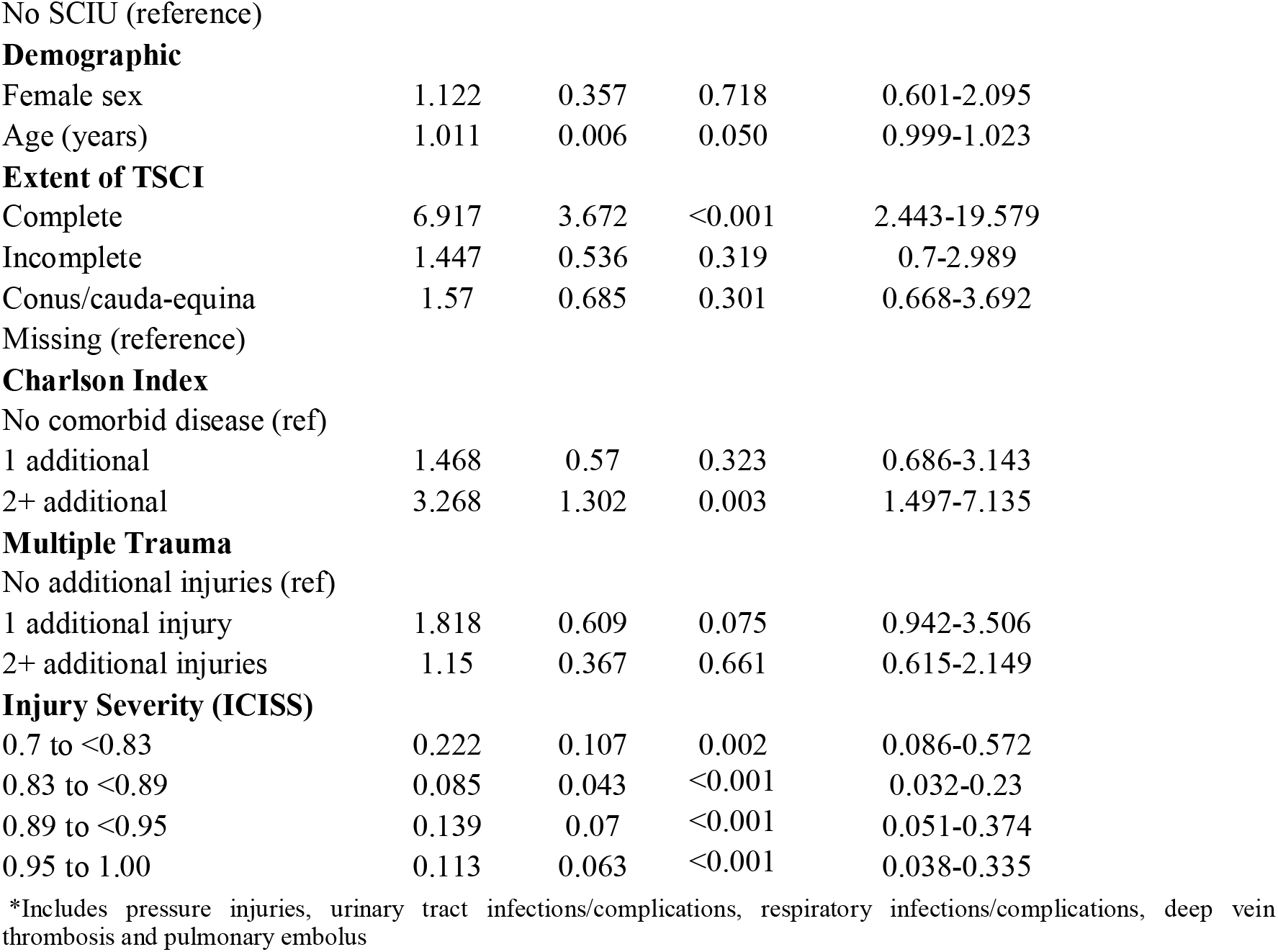
Triage pattern impact on risk of developing a medical complication as an inpatient.

| **Any complication\*** | **Odds Ratio** | **Std.Err** | **P-value** | **95% Conf. Interval** |
| --- | --- | --- | --- | --- |
| **Triage Patterns** |  |  |  |  |
| Direct SCIU | 1.392 | 0.492 | 0.35 | 0.696-2.783 |
| Indirect SCIU | 3.243 | 1.113 | 0.001 | 1.656-6.353 |
| **Demographic** |  |  |  |  |
| Female sex | 1.122 | 0.357 | 0.718 | 0.601-2.095 |
| Age (years) | 1.011 | 0.006 | 0.050 | 0.999-1.023 |
| **Extent of TSCI** |  |  |  |  |
| Complete | 6.917 | 3.672 | <0.001 | 2.443-19.579 |
| Incomplete | 1.447 | 0.536 | 0.319 | 0.7-2.989 |
| Conus/cauda-equina | 1.57 | 0.685 | 0.301 | 0.668-3.692 |
| 1 additional | 1.468 | 0.57 | 0.323 | 0.686-3.143 |
| 2+ additional | 3.268 | 1.302 | 0.003 | 1.497-7.135 |
| 1 additional injury | 1.818 | 0.609 | 0.075 | 0.942-3.506 |
| 2+ additional injuries | 1.15 | 0.367 | 0.661 | 0.615-2.149 |
| **Injury Severity (ICISS)** |  |  |  |  |
| 0.7 to <0.83 | 0.222 | 0.107 | 0.002 | 0.086-0.572 |
| 0.83 to <0.89 | 0.085 | 0.043 | <0.001 | 0.032-0.23 |
| 0.89 to <0.95 | 0.139 | 0.07 | <0.001 | 0.051-0.374 |
| 0.95 to 1.00 | 0.113 | 0.063 | <0.001 | 0.038-0.335 |

\*Includes pressure injuries, urinary tract infections/complications, respiratory infections/complications, deep vein thrombosis and pulmonary embolus

## DISCUSSION

This study investigated the impact of geospatial variables on access to specialist care for patients with acute TSCI across NSW, and revealed that despite over half of the patients being injured within a 60-minute road travel time of a SCIU, less than 30% of them experienced a direct SCIU admission. This is poor by international comparison; Cheng et al^15^ in Canada demonstrating 77% of patients with acute TSCI and within 40km of a SCIU admitted directly to a specialist unit. In this study, patients transferred directly to a SCIU underwent earlier surgery (median (IQR) of 7.3 (6; 19) hours) than patients with indirect admission to SCIU, who waited 10 hours longer to have their surgery (median (IQR) 17.5 (11; 31) hours). For many patients, indirect admission places them outside the recommended time window for surgery of ≤24 hours after injury, and at greater risk for medical complications and compromised recovery.^33^

As specialist health services, the SCIUs in NSW are located in areas of high population density, however, serve large geographical areas. Increased travel time between the accident location and SCIU reduces the likelihood of direct transfer, indicating lack of equitable access to specialist care from all areas of NSW. The fact that 70% of patients who were injured within 60 minutes of road travel to a SCIU did not experience direct transfer, suggests variations in clinical practice that leads to inequity in patient treatment. Further, almost one-fifth of patients were taken first to non-trauma designated hospitals (n=61, 19.3%), consistent with previous research showing that despite recommended pathways for patients with TSCI in NSW^25^ some acute trauma patients are still being triaged to non-trauma services.^19^

Aeromedical retrieval offers reduced travel times for the regional or very remote patient, demonstrated by these findings, however, there are resource restraints to this mode of transport. Scenario modelling could demonstrate the impact of changing transport modes to patients for whom distances by road are impediments to timely acute care. The additional resource required to retrieve patients with acute TSCI aeromedically needs to be justified by comprehensive economic evaluation. The cost per mission was estimated in 2011 at between $9,300 and $19,000;^34^ the cost benefit of such retrieval outlay over the long term costs of a TSCI has not been sufficiently explored in Australia.

Patients were less likely to experience direct transfer to SCIU when they had less severe spinal cord impairment, such as conus and cauda equina injury. This may be appropriate, however, increasing age also reduced the likelihood of direct transfer; and older patients are known to have higher risks of hospital complications and poorer outcomes with delayed intervention.^8^ As surgical intervention may not have been appropriate in some older patients, we are unable to ascertain if there had been a discussion with the SCIU regarding the appropriateness of transfer to the Specialist Unit. It would seem likely that the combination of an incomplete spinal cord injury and increasing age represents the central cord syndrome group (although this was not specifically identified in this study). This group typically present after minor falls and may not be immediately recognised as TSCI^35^, so may have missed being treated according to the transfer guidelines.

### Limitations

This study had several limitations. As we required geographic variables, we were unable to include the complete cohort of patients with acute incident TSCI identified across the study in this analysis. Therefore, a selection bias was possible, given omission of certain patients. However, patients not included were found to have similar characteristics across relevant variables (age, injury mechanism and completeness of injury), this was therefore not deemed significantly impactful on our findings.

## CONCLUSION

This study used geospatial methods to analyse pathways of care for patients with acute TSCI in NSW, who received care in a SCIU by either direct or indirect admission, and has highlighted specific areas for optimisation on a health system level. Our finding that only 30% of patients who sustained injuries within a 60-minute travel radius to a SCIU were directly admitted, is significantly lower than international comparisons and advocates improvement in early transport pathways to TSCI care.

Regardless of injury incident location, it is evident that there are transport modes and decisions can achieve early and direct SCIU admission with timely surgery. These findings complement previous research showing cost savings and reduced secondary complications achievable by streamlining care pathways^16^ for patients with TSCI. Evidence informed policy in health service optimisations can provide timely and definitive surgical treatment for those patients suffering an acute TSCI, leading to improved neurological outcomes and reduced secondary complications and mortality.

## Data Availability

Data is administrative linked and fully deidentified data and can be sought by application to the Centre for Health Record Linkage NSW, Australia

## Competing Interests

No authors declare any competing interests.

## Personal Financial Interests

No competing financial interests exist.

## Funding

LNS was funded by icare ™ for the duration of this research.

## Other Competing Interests

No other competing interests are declared by all authors.

## Notes

### Competing Interest Statement

The authors have declared no competing interest.

### Author Declarations

The NSW Population Research Ethics Committee (2012/09/420) approved this project.

